# Potential public health and economic impact of the next-generation COVID-19 vaccine mRNA-1283 in the Netherlands

**DOI:** 10.64898/2026.02.18.26346561

**Authors:** Simon van der Pol, Ekkehard Beck, Tjalke Westra, Maarten Postma, Cornelis Boersma

## Abstract

COVID-19 remains a substantial public health challenge in the Netherlands. Next-generation COVID-19 vaccine, mRNA-1283, is approved in the European Union, with potential for higher relative vaccine efficacy compared with originally-licensed COVID-19 vaccines. Its potential public health and economic impact, in adults ≥60 years and high-risk 18–59 years, was modelled versus no vaccination and originally-licensed mRNA-1273 and BNT162b2, adapting a published static Markov model with 1-year time horizon. COVID-19 burden reflected two full post-pandemic seasons. Vaccine efficacy versus mRNA-1273 was based on pivotal phase 3 NextCOVE trial data; efficacy versus BNT162b2 was derived from an indirect treatment comparison. The economically justifiable price (EJP) of mRNA-1283 versus no vaccination, and price premiums over existing vaccines, were determined at a willingness-to-pay threshold of €50,000/quality-adjusted life-year (QALY) gained. Without COVID-19 vaccination, an estimated 460,000 infections, 23,800 hospitalizations and 5,300 deaths would occur. With current coverage, mRNA-1283 was estimated to prevent 68,000 infections, 5,400 hospitalizations, and 1,200 deaths, saving 9,667 QALYs and over €66.5 million in treatment costs. The EJP was €238 versus no vaccination. Compared with mRNA-1273 and BNT162b2, mRNA-1283 was estimated to prevent additional burden (e.g., 1,309 and 1,679 hospitalizations, respectively), and was cost-effective at an incremental EJP of €62 versus mRNA-1273, and €80 versus BNT162b2. The results support continued COVID-19 vaccination to mitigate the ongoing health and societal burden of SARS-CoV-2 in the Netherlands. The comparative analyses indicate that mRNA-1283 may be associated with substantial health benefits over originally-licensed mRNA vaccines; consequently, it’s use may further improve health outcomes and economic efficiency within COVID-19 vaccination programs.

## 1. Introduction

The COVID-19 pandemic has had a profound and sustained impact on population health and patient well-being, placing unprecedented pressure on healthcare systems worldwide. Since the introduction of mRNA COVID-19 vaccines, substantial reductions in severe disease, hospitalizations, and mortality have been observed. These vaccines were developed to provide strong protection against COVID-19, and their high effectiveness has been consistently demonstrated in both clinical trials [1–3] and real-world studies [4, 5], including evidence from the Netherlands showing robust protection against severe outcomes [6].

In recent years, the Dutch Health Council has narrowed its recommendations for COVID-19 vaccination [7]. Current guidelines recommend vaccination primarily for individuals 18 years and older at increased high medical risk, adults aged ≥60 years, adults aged ≥50 years who are also eligible for seasonal influenza vaccination, and healthcare workers [7]. This policy shift reflects a declining annual burden of disease, with reductions in reported cases, hospital admissions, and COVID-19–related deaths [8].

However, COVID-19 continues to pose a substantial public health challenge in the Netherlands. Hospitalizations and mortality attributable to COVID-19 persist, in particular, among those most vulnerable to severe COVID-19 disease i.e., people with underlying medical conditions and older adults ≥60 years of age [7]. Modelling studies, as well as observational data, suggest that in the absence of vaccination, these numbers would be substantially higher [9]. The decreasing trend in vaccination coverage, from around 47% in 2024 to around 42% in 2025 [10], is also cause for concern. Moreover, COVID-19 has multiple peaks throughout the year, and the largest peak coincides with influenza and respiratory syncytial virus (RSV) circulation during the winter months [7, 11]. This temporal overlap increases pressure on healthcare capacity, underscoring the continued importance of effective preventive strategies [8].

A next-generation COVID-19 vaccine, mRNA-1283 (mNEXSPIKE, Moderna), has recently been approved in the European Union [12]. It was developed to focus the immune responses to increase protection against COVID-19 at a lower mRNA dose of 10μg (e.g., 1/5th of the dose of mRNA-1273 (Spikevax, Moderna [13]), and its innovative design allows for inclusion in respiratory combination vaccines in future further developments. In pivotal phase 3 randomized clinical trials, higher immunogenicity and a higher relative vaccine efficacy (rVE) point estimate were observed with mRNA-1283 versus mRNA-1273, with the highest differences observed in adults ≥65 years and those with underlying medical conditions [3, 14, 15]. Observational studies and GRADE meta-analyses have shown that mRNA-1273 was more effective than BNT162b2 (Comirnaty, Pfizer-BioNTech) in the real-world setting [16–20]. Building on this evidence, and an indirect treatment comparison suggesting mRNA-1283 may be more effective than BNT162b2 [21], it is essential to understand the potential added clinical and economic value of mRNA-1283 over originally licensed mRNA COVID-19 vaccines.

The objectives of this study were to assess the potential public health and economic impact of mRNA-1283 compared with no COVID-19 vaccination, considering the current endemic and seasonal COVID-19 context. Furthermore, the impact of mRNA-1283 vaccination was compared with mRNA-1273 and BNT162b2.

## 2. Materials and Methods

The public health impact and economically justifiable price (EJP) of the next-generation mRNA-1283 vaccine versus no vaccination, mRNA-1273, and BNT162b2 was estimated in the Netherlands with three different scenarios based on the post-pandemic epidemiology. A previously published static Markov decision tree model with monthly cycles comparing mRNA-1273 to no vaccination and to BNT162b2 vaccination over a 1-year time horizon in the Netherlands [9] was adapted and updated for this analysis.

### Target population

The base case target vaccination population included all adults aged ≥60 years and high-risk adults aged 18-59 years, including both the high medical risk individuals 18-49 and the flu target group 50-59 years, according to current Dutch recommendations [9].

### Model structure and inputs

The model structure and assumptions were previously described for economic analyses in the Netherlands (Zeevat et al. 2025 [9]), the United States (US) [22], and Canada [23]. In summary, individuals had a monthly risk of COVID-19 infection based on Dutch age-specific incidence data. Following infection, the risk of health outcomes was determined by the decision tree. Every month, individuals could receive a COVID-19 vaccine, based on age-specific coverage rates, which reduced the risk of infection and hospitalization following infection, according to each vaccine’s efficacy (Table 1).

**Table 1.** Vaccine efficacy of mRNA-1283, mRNA-1273 and BNT162b2 [22].

| Age group (years) | Comparison of mRNA-1273 & mRNA-1283 |  |  | Comparison of mRNA-1283 & BNT162b2 |  |  |
| --- | --- | --- | --- | --- | --- | --- |
|  | 2024/25 mRNA-1273 VE | rVE of mRNA-1283 vs mRNA-1273 (95% CI) | mRNA-1283 VE (Range) | mRNA-1283 VE | rVE of mRNA-1283 vs BNT162b2 (95% CI) | BNT162b2VE (Range) |
|  | **Symptomatic Infection** |  |  |  |  |  |
| 18–59 high-risk | 48.2% | 16.3%<br>(1.8–28.7%) | 56.6%<br>(49.1–63.0%) | 56.6% | 19.0%<br>(4.9–31.0%) | 46.5%<br>(37.2–54.4%) |
| ≥60 | 54.5% | 13.5%<br>(-7.7–30.6%) | 60.7%<br>(50.8–68.4%) | 60.7% | 22.8%<br>(3.7–38.1%) | 49.0%<br>(36.5–59.2%) |
|  | **Hospitalization<sup>†</sup>** |  |  |  |  |  |
| 18–59 high-risk | 50.5% | 38.1%<br>(-6.6–64.1%) | 69.4%<br>(46.3–82.2%) | 69.4% | 44.1%<br>(3.2–67.7%) | 45.2%<br>(5.1–68.3%) |
| ≥60 | 57.2% | 38.1%<br>(-6.6–64.1%) | 73.5%<br>(53.3–84.6%) | 73.5% | 44.1%<br>(3.2–67.7%) | 52.6%<br>(17.9–72.6%) |

<sup>†</sup> If VE against Hospitalization is lower than VE against infection, the incremental protection against the probability of hospitalization given infection is set to 0. This assumption is aligned with Fust et al. 2026 [22], and based on the observance that mRNA COVID-19 vaccines showed consistently higher protection against severe COVID-19 disease (i.e., hospitalization, death) than COVID-19 infection.
CI: Confidence interval; rVE: relative vaccine effectiveness; VE: Vaccine effectiveness; vs: versus

The model inputs and assumptions were previously described in Zeevat et al. 2025 [9]. Updates to input parameters are summarized below and in Table A1. All other input parameters follow Zeevat et al. 2025. Cost estimates from Zeevat et al. 2025 were inflated from 2023 to 2025 using the Dutch consumer price index [24].

### Model input parameter updates

As COVID-19 incidence is evolving and has seasonal fluctuations, the average incidence over two recent post-pandemic seasons was considered in the base case, as is often done for influenza modelling [25]. Thus, the base case risk of infection assumed the average incidence of symptomatic COVID-19 infections in 2023/2024 and 2024/2025 seasons in the Netherlands. The incidence for 2023/2024 was based on the incidence assumed in Zeevat et al. 2025 and reflects a high incidence scenario. Based on the recently published Netherlands infectious disease report of 2024 [8], the incidence of 2024/2025 season was estimated to be 40% of the incidence in 2023/2024 (low incidence scenario). The base case incidence, i.e. the average incidence across 2023/2024 and 2024/2025 was, thus, estimated applying a rate of 0.7 to the incidence assumed in Zeevat et al. 2025. Low and high incidence scenarios were explored in sensitivity analyses.

The vaccine coverage rate (VCR) for the Dutch campaign, running from September to December, was updated using 2025 observed values [10].

A Dutch observational study found that the risk of developing post-COVID-19 condition has declined during post-pandemic years [26]. Thus, contrary to Zeevat et al. 2025, post-COVID condition was not considered in the base case analysis as part of the infection consequences tree. However, post-COVID-19 condition was considered in a scenario analysis based on the parameters assumed in Zeevat et al. 2025 due to the substantial level of evidence on the risk of onset of post-COVID-19 condition in people infected with COVID-19, especially those most vulnerable to severe COVID-19 disease. In addition, there is an emerging level of evidence that COVID-19, like seasonal influenza and other respiratory diseases, can trigger and exacerbate chronic conditions following an COVID-19 infection [27, 28].

### Vaccine characteristics

Vaccine effectiveness (VE) inputs against COVID-19 infection and hospitalization were based on the estimates assumed in a cost-effectiveness analysis of mRNA-1283 in the US for 2025/2026 [22]. In short, the absolute VE of mRNA-1283 was estimated considering a risk ratio based approach, applying the rVE of mRNA-1283 versus mRNA-1273 (derived from the pivotal phase 3 NextCOVE trial [3]) to the VE of mRNA-1273. The latter was derived from a large real-world evidence study of mRNA-1273 VE (targeting the KP.2 variant) against COVID-19 related hospitalizations and medically-attended COVID-19 (a proxy for infections) [29]. The rVE of mRNA-1283 versus BNT162b2 was based on findings of an indirect treatment comparison [21], leading to absolute BNT162b2 VE estimates (Table 1).

In the base case, the rVE against hospitalization was based on an FDA-defined severe COVID-19 endpoint of the NextCOVE trial [3], assumed as a proxy for rVE against hospitalization [22]. The same monthly VE waning rates were assumed for all vaccines (4.75% against infection [30] and 2.46% against hospitalization [31]). Further details of the estimation of VE inputs for mRNA-1283, mRNA-1273, and BNT162b2 are presented in Fust et al. 2026 [22].

Frequencies of reactogenicity grade 3 events for mRNA-1283 and mRNA-1273 were based on the NextCOVE trial data, and grade 3 events were assumed to be the same for mRNA-1273 and BNT162b2 [23]. Rates of myocarditis/pericarditis (0.0008%) associated with all mRNA COVID-19 vaccines were assumed for high-risk adults aged 18-60 years, following estimates provided in the US FDA label [32].

Updates were made to the utility losses associated with some infection-related health states, and with adverse events [23].

### Vaccine unit costs

COVID-19 vaccine procurement in the Netherlands is conducted through European tenders and commonly includes confidential discounts, thus, net vaccine prices are not publicly disclosed. Consequently, for the economic analysis, the EJP was estimated at a willingness-to-pay (WTP) threshold of €50,000 per QALY gained, based on the proposed WTP threshold in 2024 for public health interventions including vaccines [9].

### Sensitivity and scenario analyses

The robustness of the results was assessed in sensitivity and scenario analyses. In particular, the impact of specific parameter assumptions and modelling settings was assessed on the EJP at a WTP of €50,000/QALY. The VE of mRNA-1283 against infection and hospitalization was varied using the upper and lower bounds of the 95% confidence interval; the rVE against hospitalization was varied by applying the same rVE for mRNA-1283 versus mRNA-1273 for infection to hospitalization. The VCR was also varied by ±20%, and further scenario analyses explored restricting vaccination target populations to all adults aged ≥75 years only (n=1,759,000); and of expanding to all 50-59-year-olds (n=2,465,000), instead of just high-risk 50-59-year-olds (n=298,000) [33, 34]. Lastly, a scenario analysis assessed the burden of post-COVID-19 condition and impact of vaccination. As in Zeevat et al. 2025 [9], all COVID-19 survivors were assumed to experience following the acute COVID-19 infection healthcare resource use and utility loss, while those with post-COVID-19 condition were assumed to experience productivity losses, to avoid double counting, and due to a lack of data.

Additionally, deterministic sensitivity analyses (DSA) were performed to assess the impact of varying inputs on the ICER of mRNA-1283 vaccination versus no vaccination. Following Zeevat et al. 2022 [35], the base case EJP at a WTP €50,000/QALY was considered as vaccine price for this DSA, and was varied along with other input parameters by ±20% (Zeevat et al. 2025 [9]). The DSA results are presented in a Tornado diagram.

Probabilistic sensitivity analyses (PSA) with n=1,000 simulations were conducted to assess the impact of parameter uncertainty on incremental EJPs of mRNA-1283 versus mRNA-1273 and mRNA-1283 versus BNT162b2 at a WTP threshold of €50,000/QALY following the approaches taken in Zeevat et al. 2025 [9] and Fust et al. 2025 [22] (see Supplementary materials for details on parameter input estimates). Tables A2 and A3 specify for each parameter considered in the PSA the point estimate, distribution assumed and standard error/deviation. The PSA results are presented showing the likelihood of varying incremental EJPs being cost-effective at a WTP threshold of €50,000/QALY.

## 3. Results

### 3.1. Base case analysis: mRNA-1283 versus no vaccination

With no COVID-19 vaccination in the base case, among the 14.8M adults in the Netherlands, there were an estimated 460,000 symptomatic infections, 24,000 hospitalizations and 5,500 deaths from COVID-19.

In the base case, mRNA-1283 vaccination of adults aged ≥60 years and of high-risk adults aged 50–59 years (around 2.1 million individuals vaccinated) was estimated to produce important public health gains. Around 68,000 symptomatic infections, 5,400 hospitalizations, and 1,200 deaths from COVID-19 were estimated to be prevented, with treatment cost savings exceeding €66.5 million and with 9,667 QALYs gained (Table 2). The public health impact was estimated to be between 3,000 and 7,700 hospitalizations prevented, considering a lower or higher COVID-19 incidence (Table 2).

**Table 2.** Impact of mRNA-1283 versus no vaccination.

|  | Base case |  | High incidence |  | Low incidence |  |
| --- | --- | --- | --- | --- | --- | --- |
|  | No vaccine | mRNA-1283 | No vaccine | mRNA-1283 | No vaccine | mRNA-1283 |
| Symptomatic infections | 459,068 | 391,139 | 655,697 | 558,682 | 262,371 | 223,543 |
| Averted |  | 67,929 |  | 97,015 |  | 38,828 |
| Hospitalizations | 23,830 | 18,455 | 34,037 | 26,359 | 13,620 | 10,548 |
| Averted |  | 5,375 |  | 7,677 |  | 3,072 |
| Deaths | 5,348 | 4,137 | 7,638 | 5,908 | 3,057 | 2,364 |
| Averted |  | 1,211 |  | 1,730 |  | 692 |
| Treatment costs (€) | 346,486,776 | 279,941,386 | 494,893,571 | 399,850,768 | 198,027,466 | 159,992,774 |
| Averted |  | 66,545,390 |  | 95,042,961 |  | 38,034,692 |
| QALY loss | 43,921 | 34,254 | 62,732 | 48,890 | 25,103 | 19,613 |
| QALY gain |  | 9,667 |  | 13,842 |  | 5,490 |
| EJP (WTP €50k) | €237.93 |  | €349.47 |  | €126.34 |  |

EJP: economically justifiable price; k: thousand; QALY: quality-adjusted life-year; WTP: willingness-to-pay

In the base case, the estimated number needed to vaccinate (NNV) with mRNA-1283 was 31 to prevent a symptomatic infection, 396 for a COVID-19 hospitalization, and 1,756 for a COVID-19 death.

Given a base case WTP threshold of €50,000/QALY gained, mRNA-1283 was cost-effective at an EJP of €237.93 compared with no vaccination. When considering a WTP of €20,000/QALY, the average EJP of mRNA-1283 across all three incidence scenarios was €101.57.

#### 3.1.1. Additional scenario analyses and DSA results

Varying the VE of mRNA-1283 against infection and hospitalization, using the lower bounds and upper bounds of the 95% confidence intervals, resulted in 3,754–6,693 hospitalizations prevented and 846–1,508 deaths prevented, with an EJP of €160.70 (using lower bounds) to €301.05 (using upper bounds) (Table 3). Assuming the same rVE of mRNA-1283 against infection as for hospitalization resulted in fewer prevented hospitalizations (4,532) and deaths (1,021), and a lower EJP (€199.27) versus the base case. Varying the base case VCR by ±20% resulted in 4,300–6,451 hospitalizations prevented and 969–1,453 deaths prevented but did not impact the EJP as a static health economic model was applied. Expanding the target population to all 50–59-year-olds prevented slightly more hospitalizations (5,481) and deaths (1,219) than in the base case, and lowered the EJP to €186.54 (at WTP of €50,000/QALY). By contrast, reducing the target population to only adults ≥75 years reduced the number of prevented hospitalizations (2,779) and deaths (628) considerably, and increased the EJP to €371.03 compared with the base case. The scenario assessing the impact of mRNA-1283 vaccination on post-COVID-19 condition found mRNA-1283 would prevent an estimated 2,265 incident post-COVID-19 condition cases, at an EJP of €241.96.

**Table 3.** Additional scenario analyses mRNA-1283 versus no vaccination.

|  | Hospitalizations<br>Averted | Deaths averted | EJP (WTP<br>€50,000/QALY) |
| --- | --- | --- | --- |
| **Base case** | **5,375** | **1,211** | **237.93** |
| VE infection and hospitalization<br>95% CI lower bound | 3,754 | 846 | 160.70 |
| VE infection and hospitalization<br>95% CI upper bound | 6,693 | 1,508 | 301.05 |
| rVE hospitalization = rVE infection | 4,532 | 1,021 | 199.27 |
| VCR -20% | 4,300 | 969 | 237.93 |
| VCR +20% | 6,451 | 1,453 | 237.93 |
| Expand to all 50–59-year-olds | 5,481 | 1,219 | 186.54 |
| $\geq 75$ years only | 2,779 | 628 | 371.03 |
| Post-COVID-19 condition | Additional 2,265 post-COVID-19<br>condition cases prevented |  | 241.96 |

EJP: economically justifiable price; QALY: quality-adjusted life-year; WTP: willingness-to-pay

Figure 1 shows the ten parameters with the greatest impact on the incremental cost-effectiveness ratio (ICER) of mRNA-1283 versus no vaccination (at an EJP based on WTP €50,000/QALY). The most important parameters were COVID-19 incidence, VE against hospitalization, and the hospitalization rate given infection.

**Figure 1.**
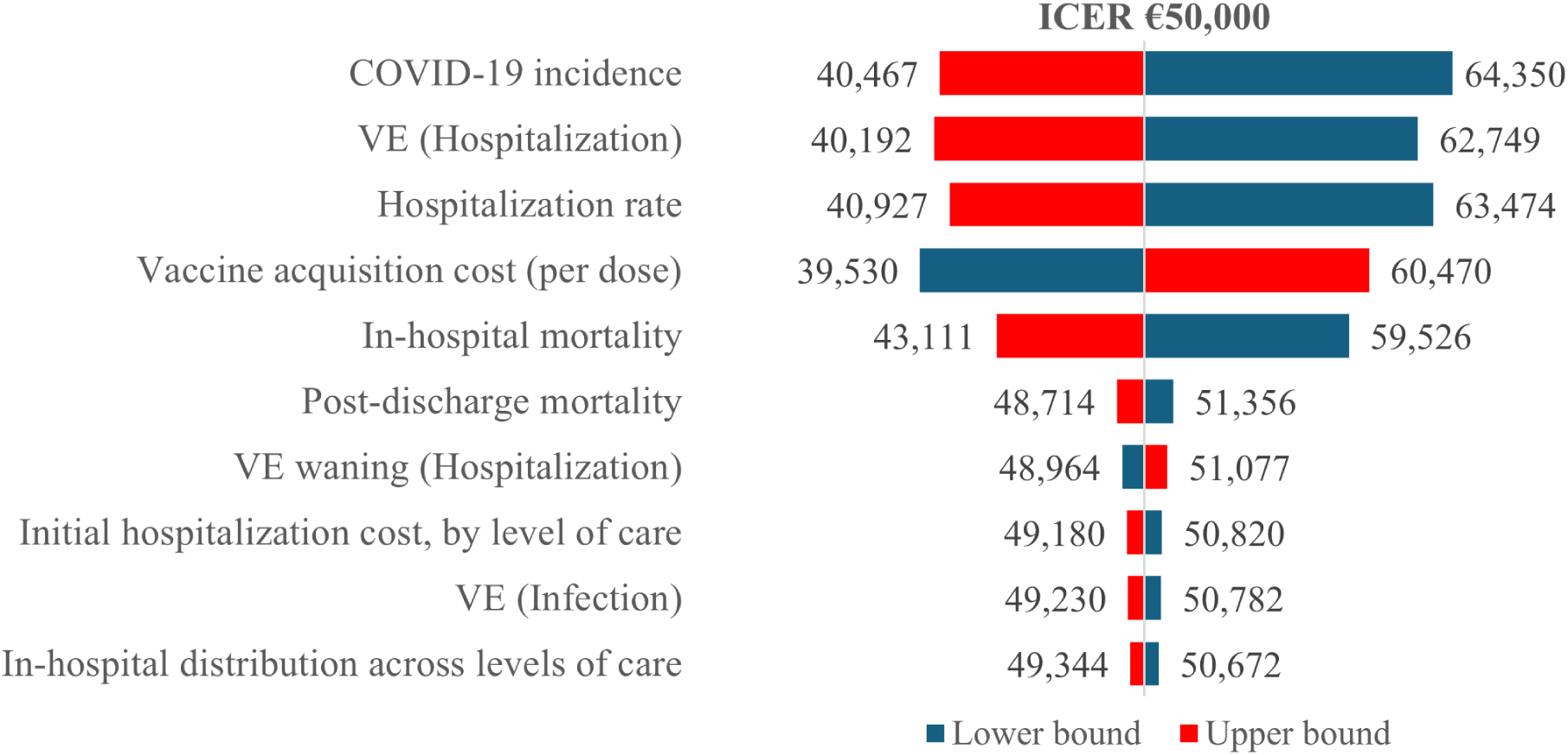
DSA results for mRNA-1283 versus no vaccination ICER: incremental cost-effectiveness ratio; VE: vaccine efficacy;

### 3.2. Analysis: mRNA-1283 versus mRNA-1273

Vaccination with mRNA-1283 prevented more symptomatic infections (8,931), hospitalizations (1,309), and COVID-19 deaths (295) than mRNA-1273, resulting in treatment cost savings (€14,743,901) and QALY gains (2,323). As expected, more vaccination gains were predicted in scenarios with a higher COVID-19 incidence and higher mRNA-1283 versus mRNA-1273 rVE (Table 4).

**Table 4.** Impact of mRNA-1283 versus mRNA-1273.

|  | Base case |  | High incidence |  | Low incidence |  | Lower bound rVE |  | Upper bound rVE |  |
| --- | --- | --- | --- | --- | --- | --- | --- | --- | --- | --- |
|  | mRNA-1273 | mRNA-1283 | mRNA-1273 | mRNA-1283 | mRNA-1273 | mRNA-1283 | mRNA-1273 | mRNA-1283 | mRNA-1273 | mRNA-1283 |
| Symptomatic infections | 400,070 | 391,139 | 571,437 | 558,682 | 228,648 | 223,543 | 400,070 | 405,036 | 400,070 | 380,290 |
| Averted | 8,931 |  | 12,755 |  | 5,105 |  | -4,966 |  | 19,780 |  |
| Hospitalizations | 19,764 | 18,455 | 28,230 | 26,359 | 11,296 | 10,548 | 19,764 | 20,077 | 19,764 | 17,563 |
| Averted | 1,309 |  | 1,871 |  | 748 |  | -312 |  | 2,201 |  |
| Deaths | 4,432 | 4,137 | 6,330 | 5,908 | 2,533 | 2,364 | 4,432 | 4,502 | 4,432 | 3,936 |
| Averted | 295 |  | 422 |  | 169 |  | -70 |  | 496 |  |
| Treatment costs | 294,856,989 | 280,113,088 | 421,155,221 | 400,096,059 | 168,517,334 | 160,090,990 | 294,856,989 | 298,821,108 | 294,856,989 | 269,157,438 |
| Averted | 14,743,901 |  | 21,059,162 |  | 8,426,344 |  | -3,964,119 |  | 25,699,552 |  |
| QALY loss | 36,571 | 34,248 | 52,206 | 45,787 | 20,930 | 19,609 | 36,571 | 37,159 | 36,571 | 32,633 |
| QALY gain | 2,323 |  | 3,325 |  | 1,321 |  | -588 |  | 3,938 |  |
| Incremental EJP (WTP 50k) | €61.53 |  | €88.06 |  | €35.00 |  | NA |  | €104.65 |  |
| Incremental EJP (WTP 20k) | €28.77 |  | €41.16 |  | €16.37 |  | NA |  | €49.11 |  |

EJP: economically justifiable price; k: thousand; NA: indicates negative incremental EJP of mRNA-1283 over mRNA-1273; QALY: quality-adjusted life-year; rVE: relative vaccine efficacy; WTP: willingness-to-pay

Using the base case incidence and rVE, mRNA-1283 was cost-effective at an incremental EJP of €61.53 over mRNA-1273 (WTP €50,000/QALY) and €28.77 (WTP €20,000/QALY). These values increased as vaccination benefits increased, in the higher incidence and higher rVE scenarios (Table 4).

In the base case, the incremental NNV, i.e. the additional number of persons to be vaccinated with mRNA-1273 versus mRNA-1283 to prevent one COVID-19 hospitalization, was 127.

With an incremental EJP of €61.53, the probability of mRNA-1283 versus mRNA-1273 being cost-effective at a WTP of €50,000/QALY was 0.44 (Figure 2).

**Figure 2.**
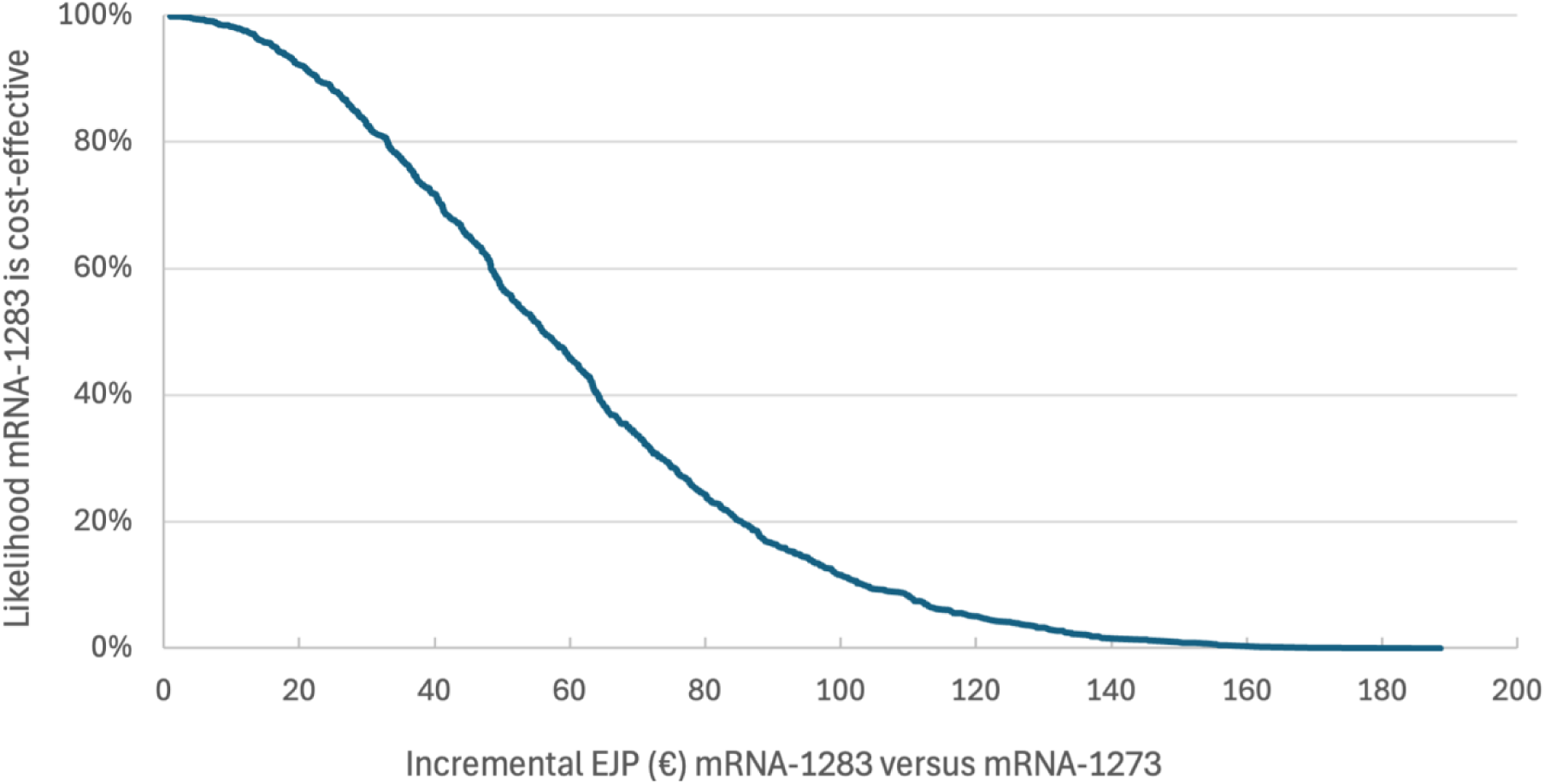
Cost-effectiveness acceptability curve for mRNA-1283 versus mRNA-1273.

### 3.3. Analysis: mRNA-1283 versus BNT162b2

Vaccination with mRNA-1283 prevented more symptomatic infections (16,499), hospitalizations (1,679), and COVID-19 deaths (378) than BNT162b2, resulting in treatment cost savings (€19,827,071) and QALY gains (3,005). Again, more vaccination gains were predicted in scenarios with a higher COVID-19 incidence and higher rVE (Table 5).

**Table 5.**
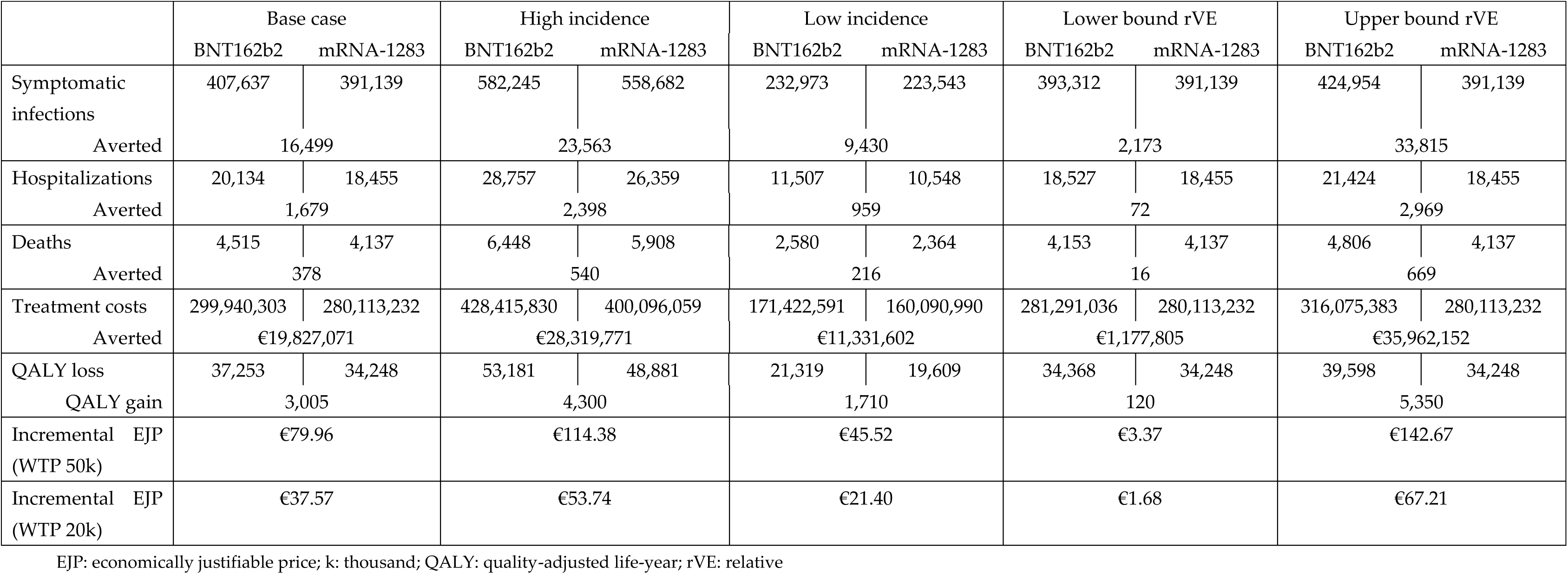
Impact of mRNA-1283 versus BNT162b2.

|  | Base case |  | High incidence |  | Low incidence |  | Lower bound rVE |  | Upper bound rVE |  |
| --- | --- | --- | --- | --- | --- | --- | --- | --- | --- | --- |
|  | BNT162b2 | mRNA-1283 | BNT162b2 | mRNA-1283 | BNT162b2 | mRNA-1283 | BNT162b2 | mRNA-1283 | BNT162b2 | mRNA-1283 |
| Symptomatic infections | 407,637 | 391,139 | 582,245 | 558,682 | 232,973 | 223,543 | 393,312 | 391,139 | 424,954 | 391,139 |
| Averted | 16,499 |  | 23,563 |  | 9,430 |  | 2,173 |  | 33,815 |  |
| Hospitalizations | 20,134 | 18,455 | 28,757 | 26,359 | 11,507 | 10,548 | 18,527 | 18,455 | 21,424 | 18,455 |
| Averted | 1,679 |  | 2,398 |  | 959 |  | 72 |  | 2,969 |  |
| Deaths | 4,515 | 4,137 | 6,448 | 5,908 | 2,580 | 2,364 | 4,153 | 4,137 | 4,806 | 4,137 |
| Averted | 378 |  | 540 |  | 216 |  | 16 |  | 669 |  |
| Treatment costs | 299,940,303 | 280,113,232 | 428,415,830 | 400,096,059 | 171,422,591 | 160,090,990 | 281,291,036 | 280,113,232 | 316,075,383 | 280,113,232 |
| Averted | €19,827,071 |  | €28,319,771 |  | €11,331,602 |  | €1,177,805 |  | €35,962,152 |  |
| QALY loss | 37,253 | 34,248 | 53,181 | 48,881 | 21,319 | 19,609 | 34,368 | 34,248 | 39,598 | 34,248 |
| QALY gain | 3,005 |  | 4,300 |  | 1,710 |  | 120 |  | 5,350 |  |
| Incremental EJP (WTP 50k) | €79.96 |  | €114.38 |  | €45.52 |  | €3.37 |  | €142.67 |  |
| Incremental EJP (WTP 20k) | €37.57 |  | €53.74 |  | €21.40 |  | €1.68 |  | €67.21 |  |

EJP: economically justifiable price; k: thousand; QALY: quality-adjusted life-year; rVE: relative

Using the base case incidence and rVE, mRNA-1283 was cost-effective at an incremental EJP of €79.96 over BNT162b2 (WTP €50,000/QALY) and €37.57 (WTP €20,000/QALY). These values increased as vaccination benefits increased, in the higher incidence and higher mRNA-1283 versus BNT162b2 rVE scenarios (Table 5).

In the base case, the incremental NNV, i.e. the additional number of persons to be vaccinated with BNT162b2 versus mRNA-1283 to prevent one COVID-19 hospitalization, was 180.

With an incremental EJP of €79.96, the probability of mRNA-1283 versus BNT162b2 being cost-effective at a WTP of €50,000/QALY was 0.45 (Figure 3).

**Figure 3.**
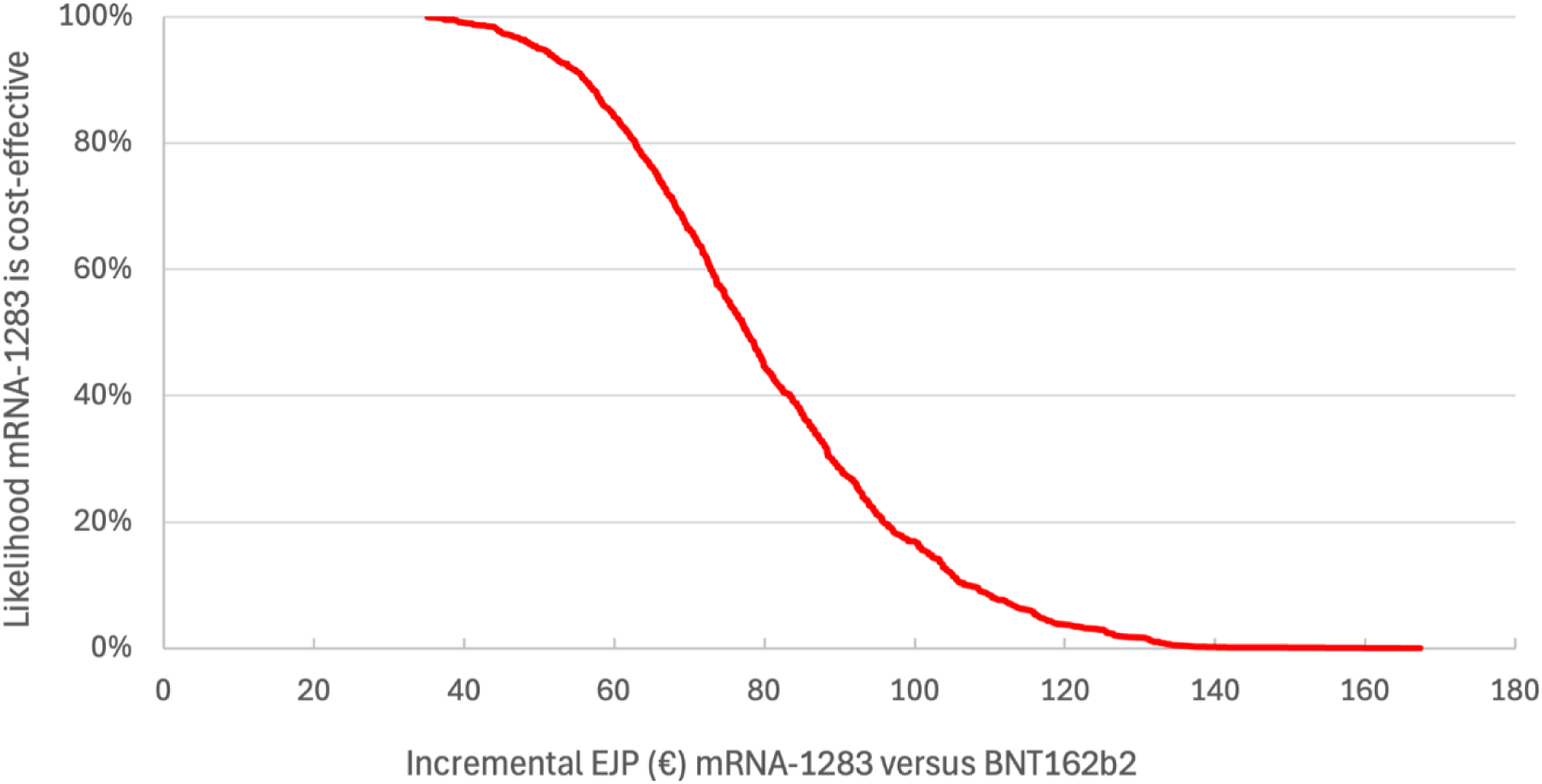
Cost-effectiveness acceptability curve for mRNA-1283 versus BNT162b2.

## 4. Discussion

In the Netherlands, the burden due to COVID-19 remains substantial, especially in those most vulnerable to severe COVID-19 disease. Our results estimated that without vaccination, 460,000 (260,000–660,000) individuals would develop symptomatic COVID-19, leading to 24,000 (13,000–34,000) hospitalizations and 5,400 (3,000–8,000) deaths. Considering the current vaccination coverage and recommended target population, a potential vaccination campaign with mRNA-1283 could prevent 68,000 (39,000–97,000) symptomatic cases, 5,400 (3,000–8,000) hospitalizations and 1,200 (700– 1,700) deaths compared with no vaccination. These health gains were estimated to lead to 9,667 QALYs gained, as well as €66.5 million in cost-savings. mRNA-1283 vaccination was estimated to be cost-effective at an EJP of €238 (WTP of €50,000/QALY) and of €102 (WTP of €20,000/QALY).

These model-based estimates, as well as reported surveillance data, suggest that despite widespread population immunity, COVID-19 continues to place a substantial burden on individuals and public health in the Netherlands under current incidence levels. Ongoing transmission results in a large number of symptomatic infections, of which approximately 30% seek care from a general practitioner, which can contribute to sustained pressure on primary care services [36]. Importantly, COVID-19 now circulates concurrently with other major respiratory pathogens, including RSV and influenza. The overlapping seasonality of these infections creates a risk of simultaneous epidemic peaks, increasing the likelihood of a so-called “triple endemic” scenario, and increasing the strain on both primary and secondary healthcare capacity. Assuming a COVID-19 hospital length of stay of 8.43 days [37], vaccination with mRNA-1283 could potentially prevent 45,000 hospital bed-days and approximately 17,000 general practitioner visits. Beyond direct healthcare utilization, COVID-19 also imposes a considerable societal burden through productivity losses due to absenteeism and reduced work capacity. Taken together, these factors underline that the current COVID-19 vaccination program remains highly relevant, from a public health perspective and from a broader societal standpoint. Maintaining protection against COVID-19 in the working-age population is essential to safeguard workforce availability and ensure the continuity of critical sectors such as healthcare, education and other essential public services.

Based on pivotal clinical trial data and findings of an indirect treatment comparison, our results estimated that mRNA1283 could provide greater public health benefits than currently available COVID-19 mRNA vaccines, especially in those most vulnerable to severe COVID-19 disease. Compared with mRNA-1273, vaccination with next generation mRNA-1283 was estimated to prevent 8,900 additional symptomatic COVID-19 infections, 1,300 additional hospitalizations, and 295 additional deaths, leading to higher healthcare cost-savings and QALY gains. Even greater health gains and cost-savings were estimated for mRNA-1283 versus BNT162b2, the currently used vaccine in the Netherlands [9] (i.e., preventing around 16,500 additional symptomatic COVID-19 infections, 1,700 additional hospitalizations, and 380 additional deaths). Potential vaccination with mRNA-1283 was cost-effective versus mRNA-1273 at an incremental EJP of €62, and versus BNT162b2 at an incremental EJP of €80, considering a WTP of €50,000/QALY for both comparisons. These findings are in line with other public health and economic evaluations comparing mRNA-1283 vaccination with no vaccination and with existing COVID-19 vaccinations in Canada [23] and the US [22], and with the previous Dutch evaluation showing the vaccination gains of mRNA-1273 over BNT162b2 [9].

This analysis estimated an EJP of €238 for COVID-19 vaccination. Compared with other preventive interventions for respiratory infectious diseases, such as influenza and RSV vaccination, the COVID-19 EJP seems to be notably higher [9, 38, 39]. This difference can largely be explained by the substantially higher burden of disease associated with COVID-19 [40, 41]. Even in the post-pandemic period, COVID-19 continues to result in a structurally higher number of hospitalizations and deaths than influenza and RSV [40]. As these parameters are key drivers of cost-effectiveness outcomes, these factors directly contribute to the higher health gains and cost offsets achievable with COVID-19 vaccination, and consequently to the higher EJP. Differences in methodological choices across evaluations further contribute to the observed variation in EJPs between respiratory vaccines [9, 38, 39]. In most recent cost-effectiveness analyses of high-dose influenza vaccination, comparisons were typically made against standard-dose influenza vaccination, capturing only the incremental benefits of switching between vaccine types, rather than the full value of influenza vaccination versus no vaccination [38]. As a result, these analyses typically do not aim to assess the overall value of influenza vaccination programs when compared with evaluations assessing vaccination versus no vaccination, as this COVID-19 analysis has done. As such, the higher EJP estimated for COVID-19 vaccination appears consistent with both the underlying epidemiology and the analytical framework applied, and reflects the persistently higher disease burden of COVID-19 relative to other respiratory infections.

Previously discussed limitations [9, 22, 23, 42] also apply to this evaluation. Uncertainty around the evolving incidence of COVID-19 was addressed by evaluating vaccination impact under low and high incidence scenarios; however, the true trajectory of COVID-19 incidence will only become clear over time. The number of COVID-19 hospitalizations reported in the Netherlands (average of 2023 and 2024, 19,500 hospitalizations [8, 43]) was comparable to the number predicted in the base case model (average of 2023/2024 and 2024/2025, range 18,455 with mRNA1283 to with 20,134 BNT162b2), providing support for the base case incidence assumptions. The efficacy and safety of mRNA-1283 has not yet been established in the real-world setting. Thus, the rVE of mRNA-1283 versus mRNA-1273 from the NextCOVE trial [3], as well as the results of the indirect treatment comparison of mRNA-1273 and BNT162b2, should be confirmed in real-world evidence studies. No data for the Netherlands (or Europe) were identified on utility loss associated with COVID-19, thus, US data were used [9].

There remains a clear need to continue investing in COVID-19 vaccination in the Netherlands. Such programs are still highly relevant given the ongoing burden of disease and additionally, new COVID-19 vaccines such as the next-generation mRNA-1283 may provide an opportunity to optimize COVID-19 vaccination programs, especially for populations most vulnerable to severe COVID-19. Although overall incidence has declined since the peak of the pandemic, COVID-19 transmission remains unpredictable, with a persistent risk of renewed increases in incidence. In addition, while vaccination of all adults aged 50–59 years may be perceived as less critical from a public health perspective, it is highly relevant from a broader societal perspective, as preventing COVID-19 and other respiratory infectious diseases in this age group helps reduce productivity losses and work absenteeism, thus providing wider socioeconomic gains.

## 5. Conclusions

The findings support continued COVID-19 vaccination as a key public health measure to address the persistent health and societal burden associated with ongoing transmission of SARS-CoV-2. In this setting, optimizing vaccine choice within existing programs becomes increasingly important. The comparative analyses suggest that mRNA-1283 is potentially associated with the greatest health gains, based on evidence of potential higher efficacy or effectiveness compared with originally-licensed mRNA COVID-19 vaccines. Consequently, using mRNA-1283 could further enhance both health outcomes and economic benefits within COVID-19 vaccination programs.

## Data Availability

All data produced in the present work are contained in the manuscript

## Author Contributions

All authors were involved in study concept and design. EB and TW parameterized and programmed the model. All authors were involved in analyses and interpretation of the data. All authors reviewed the paper critically for intellectual content. All authors approve of this version to be published and agree to be accountable for all aspects of the work.

## Institutional Review Board Statement

Not applicable.

## Informed Consent Statement

Not applicable

## Data Availability Statement

The data presented in this study are available in this article and supplementary file.

## Acknowledgments

Medical writing and editorial assistance was provided by Kavi Littlewood (Littlewood Writing Solutions) in accordance with Good Publication Practice (GPP 2022) guidelines, funded by Moderna, Inc., and under the direction of the authors.

## Abbreviations

The following abbreviations are used in this manuscript:

BNT162b2: Comirnaty, Pfizer-BioNTech
CI: Confidence interval
DSA: Deterministic sensitivity analysis
EJP: Economically justifiable price
ICER: Incremental cost-effectiveness ratio
K: thousands
M: million
mRNA-1273: Spikevax, Moderna
mRNA-1283: mNEXSPIKE, Moderna
NNV: Number needed to vaccinate
PSA: Probabilistic sensitivity analysis
QALY: Quality-adjusted life-year
rVE: Relative vaccine effectiveness
US: United States
VCR: Vaccine coverage rate
VE: Vaccine effectiveness
WTP: Willingness-to-pay

## Appendix A

**Table A1.**
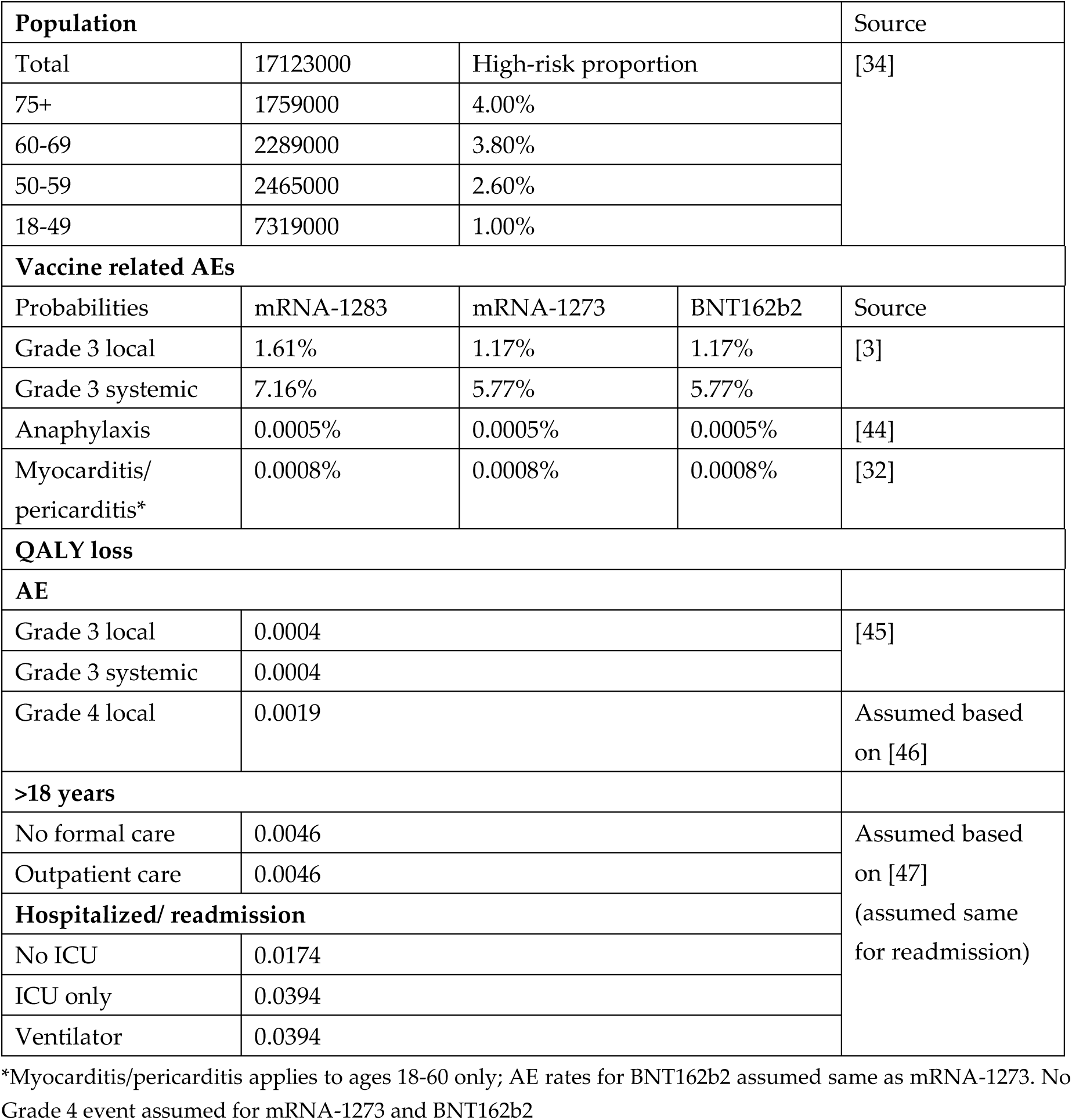
Additional updated model inputs.

**Table A2.**
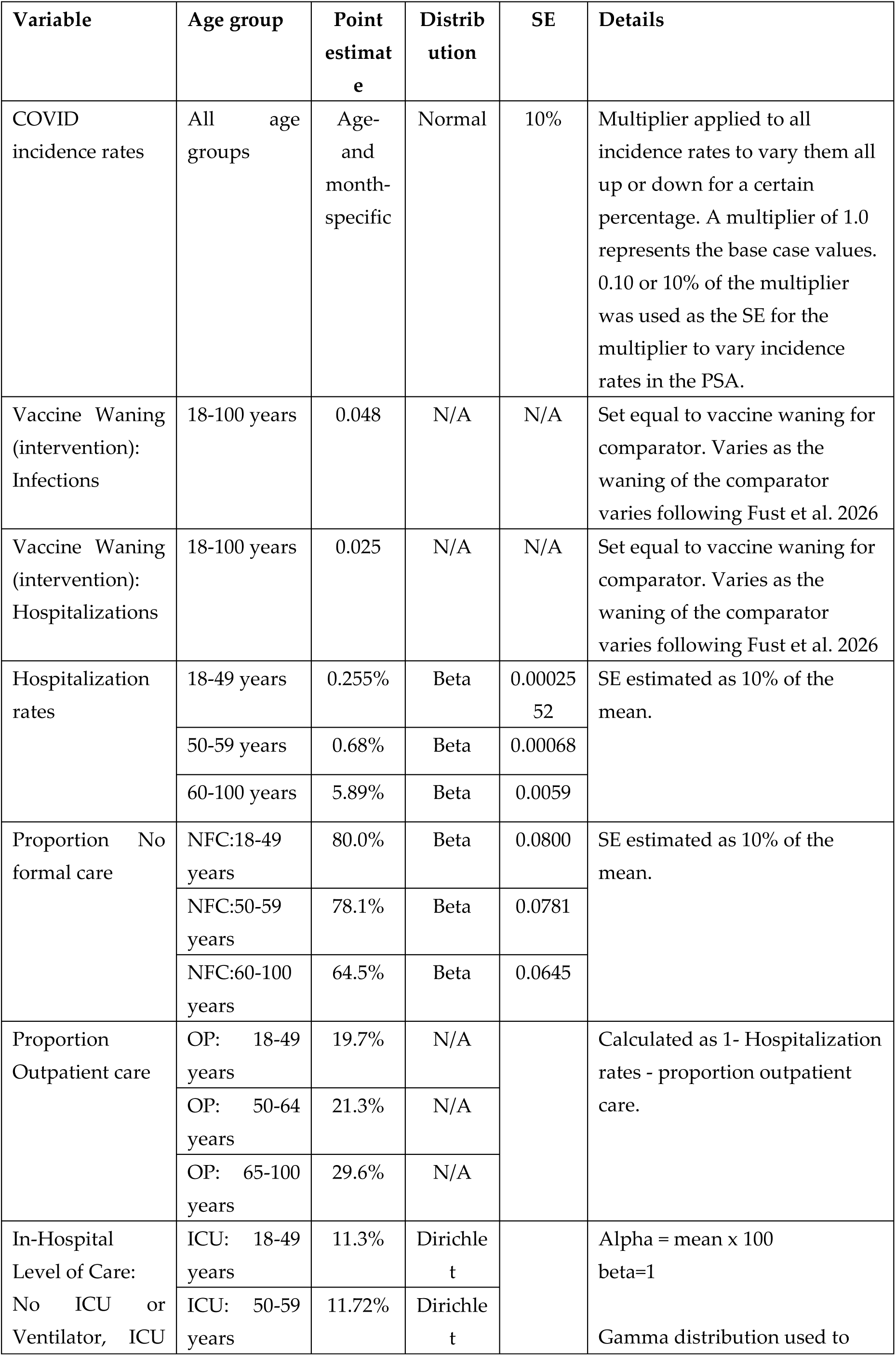
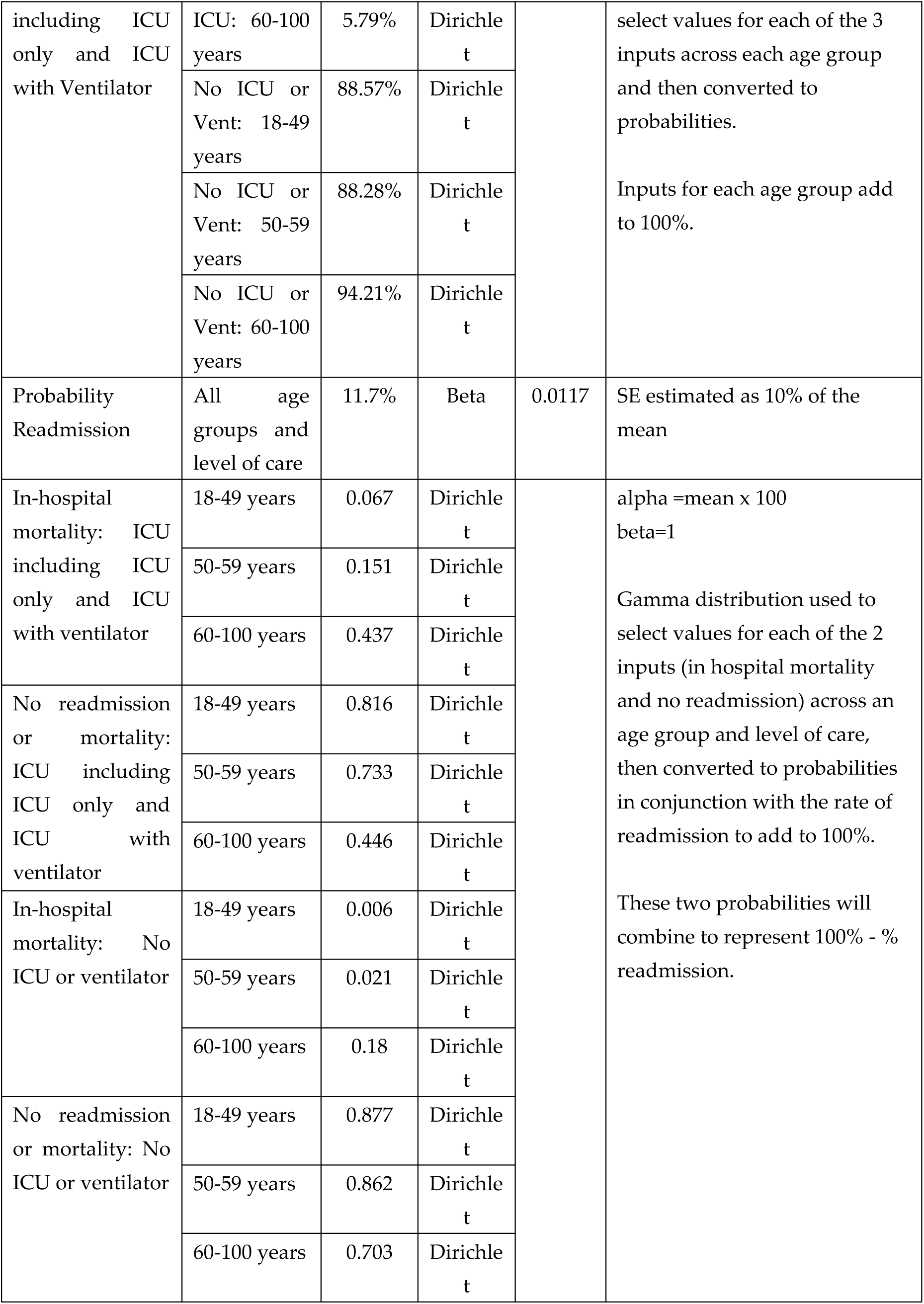
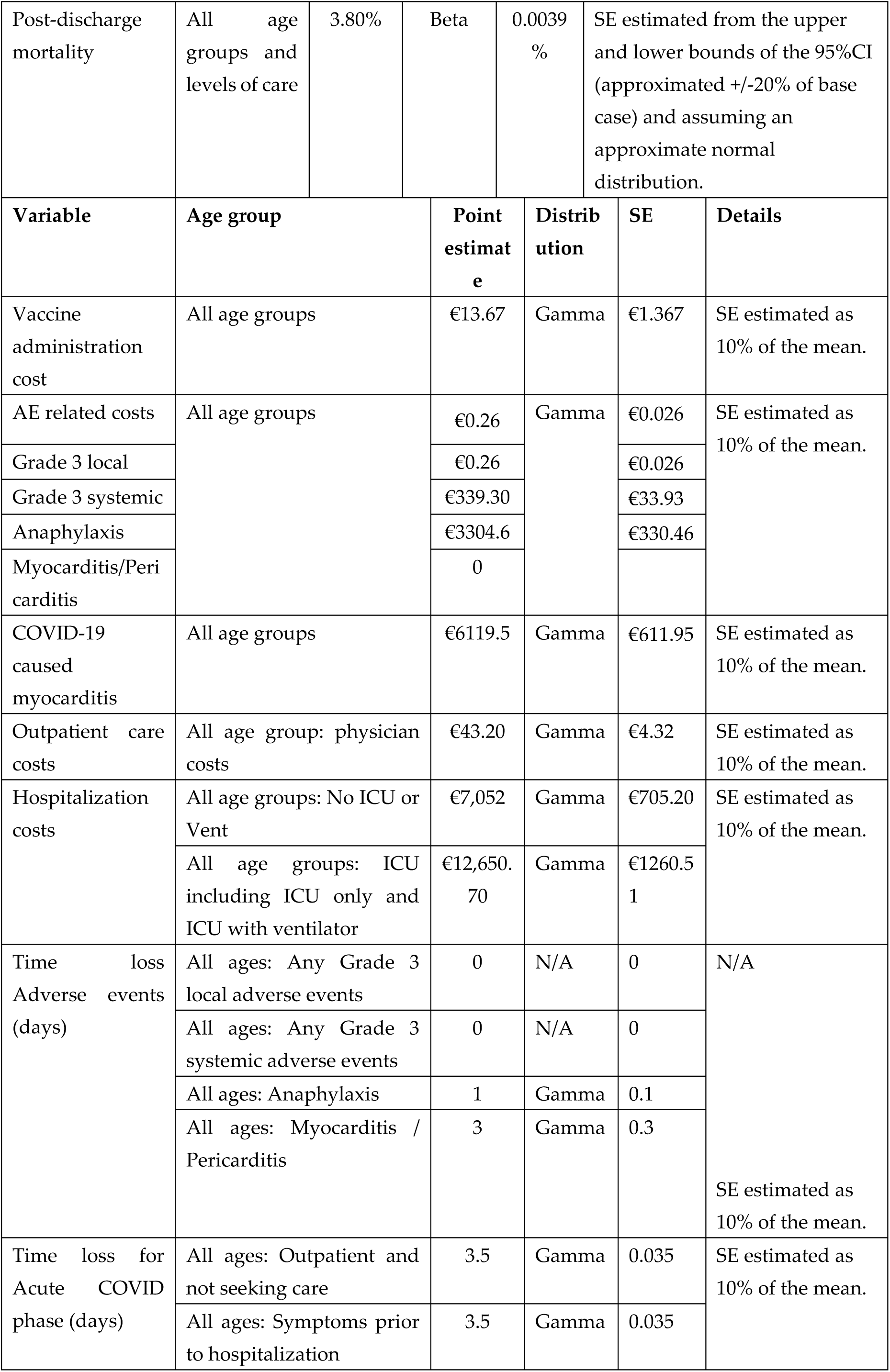
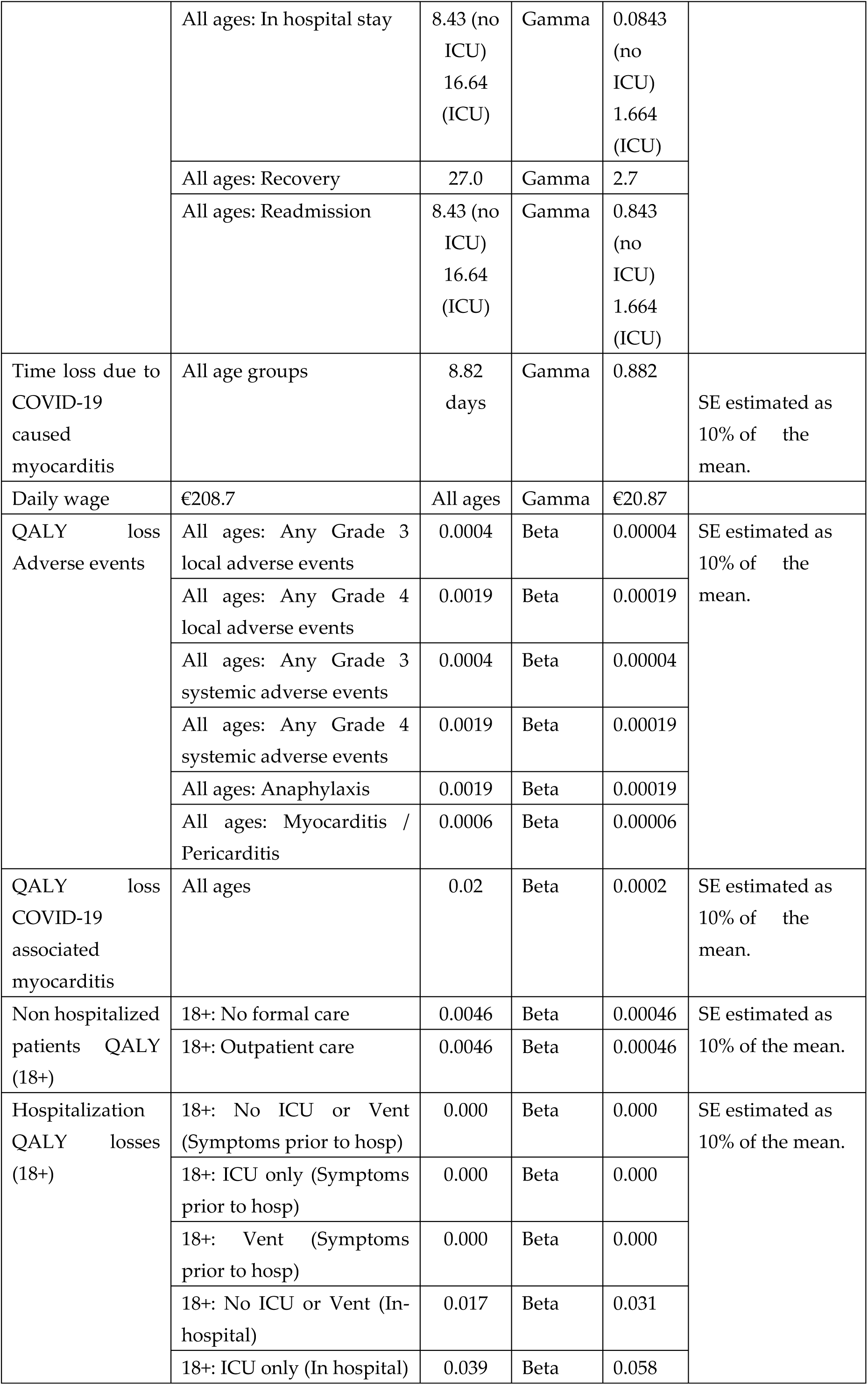
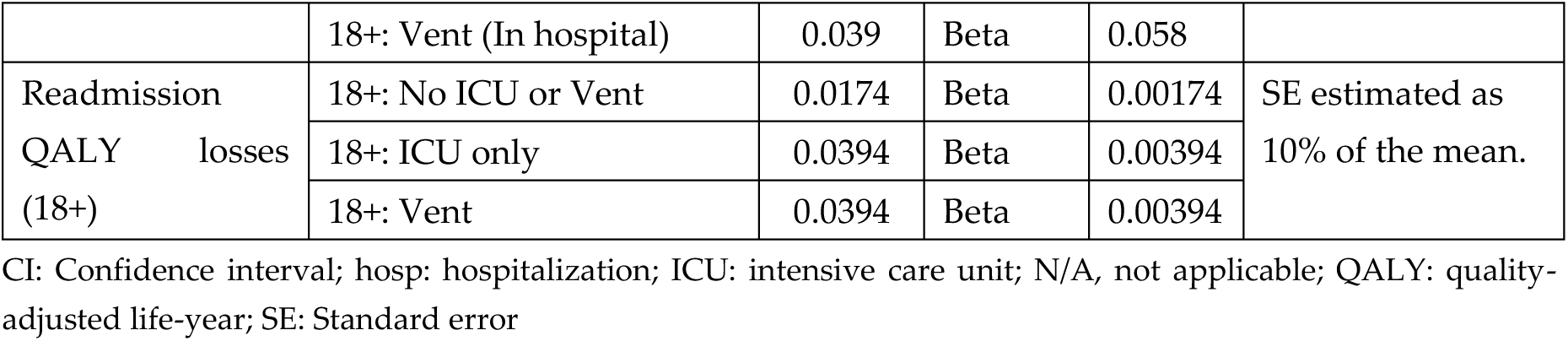
Probabilistic sensitivity analyses parameters used for comparisons of mRNA-1283 versus mRNA-1273 and mRNA-1283 versus BNT162b2.

**Table A3.** Probabilistic sensitivity analyses parameters for vaccine effectiveness and vaccine acquisition cost for mRNA-1283 versus mRNA-1273 and mRNA-1283 versus BNT162b2.

| Scenario: mRNA-1273 versus mRNA-1283 |  |  |  |  |  |
| --- | --- | --- | --- | --- | --- |
| rVE:<br>Infections | 18-49 years | 0.163 | Beta | 0.0686 | SE estimated from the upper and lower bounds of the 95%CI (including negative estimates for LB) and assuming an approximate normal distribution. |
|  | 50-59 years | 0.163 | Beta | 0.0686 |  |
|  | 60-100 years | 0.135 | Beta | 0.0977 |  |
| rVE:<br>Hospitalizations | 18-49 years | 0.381 | Beta | 0.1804 | SE estimated from the upper and lower bounds of the 95%CI (including negative estimates for LB) and assuming an approximate normal distribution. |
|  | 50-59 years | 0.381 | Beta | 0.1804 |  |
|  | 60-100 years | 0.381 | Beta | 0.1804 |  |
| 1283 Vaccine Effectiveness: Infections | 18-49 years | 0.566 | N/A | N/A | Calculated based on comparator (i.e., mRNA-1273) VE and rVE selection. Varies as other values vary. Maximum value of 0.99 included in calculations. |
|  | 50-59 years | 0.566 |  |  |  |
|  | 60-100 years | 0.607 |  |  |  |
| 1283 Vaccine Effectiveness: Hospitalizations | 18-49 years | 0.694 | N/A | N/A | Calculated based on comparator (i.e., mRNA-1273) VE and rVE selection. Varies as other values vary. Maximum value of 0.99 included in calculations. |
|  | 50-59 years | 0.694 |  |  |  |
|  | 60-100 years | 0.735 |  |  |  |
| Scenario: mRNA-1283 versus BNT162b2 |  |  |  |  |  |
| rVE:<br>Infections | 18-49 years | 0.190 | Beta | 0.0190 | SE estimated as 10% of the mean. |
|  | 50-59 years | 0.190 | Beta | 0.0190 |  |
|  | 60-100 years | 0.228 | Beta | 0.0228 |  |
| rVE:<br>Hospitalizations | 18-49 years | 0.441 | Beta | 0.0441 | SE estimated as 10% of the mean. |
|  | 50-59 years | 0.441 | Beta | 0.0441 |  |
|  | 60-100 years | 0.441 | Beta | 0.0441 |  |
| BNT162b2 Vaccine Effectiveness: Infections | 18-49 years | 0.465 | N/A | N/A | Calculated based on comparator (i.e., mRNA-1283) VE and rVE selection. Varies as other values vary. Maximum value of 0.99 included in calculations. |
|  | 50-59 years | 0.465 |  |  |  |
|  | 60-100 years | 0.490 |  |  |  |
| BNT162b2 Vaccine Effectiveness: Hospitalizations | 18-49 years | 0.452 | N/A | N/A | Calculated based on comparator (i.e., mRNA-1283) VE and rVE selection. Varies as other values vary. Maximum value of 0.99 included in calculations. |
|  | 50-59 years | 0.452 |  |  |  |
|  | 60-100 years | 0.526 |  |  |  |

CI: Confidence interval; N/A, not applicable; SE: Standard error

## Disclaimer/Publisher’s Note

The statements, opinions and data contained in all publications are solely those of the individual author(s) and contributor(s) and not of MDPI and/or the editor(s). MDPI and/or the editor(s) disclaim responsibility for any injury to people or property resulting from any ideas, methods, instructions or products referred to in the content.

